# Multi-model LLM assessment of Quality Control Circle methodological quality: a designed-anchor reliability study

**DOI:** 10.64898/2026.08.12.26360276

**Authors:** Hsuan-Ming Lin, Jr Jung Lyu

## Abstract

**Background:** Quality Control Circle (QCC) reports are often reviewed qualitatively, but reviewer workload and inter-rater variability make large-scale assessment difficult. We evaluated whether multiple large language models (LLMs) could score QCC methodological quality reliably on a designed-anchor benchmark.

**Objective:** To estimate inter-model reliability for QCC quality scoring and to assess alignment with designed anchors and descriptive comparability to a small set of public PMC QCC reports.

**Methods:** Thirty synthetic QCC reports and eight public PMC QCC reports were evaluated across four primary LLM evaluators (GPT, Gemini, Grok, DeepSeek) and one sensitivity evaluator (Claude); Claude was excluded from the primary panel because a same-family model generated the synthetic cases. Evaluators received the report text only; the trap manifest and designed-anchor scores were withheld, and their absence was verified programmatically. Each case was scored on eight QCC quality dimensions in three runs per evaluator, summarized by median, and ICC(A,1) was estimated across the primary panel. Anchor calibration, trap detection from the evaluators’ structured defect output under a map fixed before scoring, sensitivity analyses, and a descriptive synthetic-versus-PMC check were also examined.

**Results:** Inter-model reliability on the primary *k*=4 panel was good: ICC(A,1) = 0.843 (95% CI 0.812 to 0.870; case-level bootstrap 0.782 to 0.874) on 239 pooled case-dimension rows from all 30 cases; per-dimension estimates ranged from 0.747 to 0.901. The pre-specified *k*=5 analysis including Claude gave 0.833, leave-one-out estimates ranged from 0.835 to 0.857, and the originally specified score parser gave 0.854. On the 52 trap-affected case-dimension cells the primary-panel consensus fell within *±*1 of the designed anchor in 50 (96.2%), with mean absolute error 0.54 and exact agreement 34.6%; 51.5% of individual primary-panel run scores equalled the anchor. A defect of the expected category was emitted in 87% of evaluator runs and by 94% of (trap, evaluator) pairs, versus 55% and 68% for a pre-specified keyword detector; because some categories are emitted at high base rates regardless of case, these are upper bounds. All eight dimensions met the descriptive synthetic-versus-PMC margin; synthetic cases tended to score higher, significantly so only for Action Alignment.

**Conclusions:** Multi-model LLM scoring of QCC methodological quality achieved good inter-model reliability and directionally consistent alignment with synthetic anchor scores, with planted weaknesses identified far more often in structured defect output than by keyword matching. Benchmark feasibility is established; expert-labeled validation in clinical QCC reports is the planned next step.

## Introduction

Quality Control Circle (QCC) is a structured team-based quality improvement approach widely adopted in health care, particularly across East and Southeast Asian hospital systems [1, 2, 3]. A circle is a small front-line team that works through a fixed sequence of steps—topic selection, current-state survey, target setting, cause analysis (a fishbone diagram followed by verification of the “true causes”), countermeasures, effect confirmation and standardisation— and documents the cycle as a formal report built on Pareto charts, a target-setting formula and a radar chart of “intangible results”. Hospitals use these reports for internal review, quality-award competitions and accreditation evidence [4], yet their methodological quality is rarely assessed systematically.

Manual expert review is resource-intensive in the authors’ experience of hospital and competition review: reviewers must evaluate several methodological dimensions at once, and reviewer availability constrains the scale at which timely, structured feedback can be provided, which motivates interest in computational approaches that could augment review.

Large language models (LLMs) can extract structured information from clinical documents and perform evaluation tasks that require natural-language reasoning [5, 6, 7, 8], and prompt engineering can improve output consistency across model settings [9]. The precondition for any quality assessment application, however, is inter-evaluator reliability—whether a multi-LLM scoring system produces consistent results across independent evaluators and repeated runs— and this has not been addressed for QCC methodological quality scoring.

We report a designed-anchor benchmark study in which five LLM evaluators independently scored 30 synthetic Traditional-Chinese QCC reports, written in the Taiwanese hospital report format, across eight quality dimensions. The primary question was inter-model reliability, measured as ICC(A,1). Secondary analyses addressed anchor calibration, detection of planted weaknesses in the evaluators’ structured defect output, evaluator-panel sensitivity, and descriptive plausibility relative to a small set of public PMC QCC reports. The study is framed as a reliability and feasibility study; the designed-anchor design supports reliability inference but not clinical validity claims.

## Methods

### Study design and data sources

This was a cross-sectional benchmark study of LLM-based QCC quality scoring using synthetic reports; no patient data were used and no ethics review was required.

The benchmark comprised 30 synthetic QCC reports with planted trap manifests and designed-anchor quality scores at three requested quality levels (10 LOW, 14 MEDIUM, 6 HIGH). Each case embedded methodological weaknesses (traps) from a catalogue of ten trap types (Q01–Q10) that specifies, for each trap, the report section in which it is planted and which of the eight quality dimensions (Q1–Q8, named in Table 1) it degrades. The 58 planted traps fall on five dimensions (Q3, Q4, Q5, Q6, Q8; 32 of 65 trap-dimension pairs on Q5); Q1, Q2 and Q7 carry no trap and are assessed only through their anchors.

**Table 1.** Inter-model ICC(A,1) by QCC quality dimension on the primary *k*=4 panel (GPT, Gemini, Grok, DeepSeek; complete-case synthetic QCC cases). The *k*=5 sensitivity analysis adding Claude is reported in Supplementary Table S1.

| Code | Dimension | ICC(A,1) | 95% CI | Interpretation | $n$ |
| --- | --- | --- | --- | --- | --- |
| Q1 | Root Cause Depth | 0.819 | 0.694–0.903 | Good | 30 |
| Q2 | Systemic Coverage | 0.784 | 0.664–0.878 | Good | 30 |
| Q3 | Action Alignment | 0.813 | 0.676–0.901 | Good | 30 |
| Q4 | Data Foundation | 0.854 | 0.763–0.920 | Good | 30 |
| Q5 | Methodological Rigor | 0.747 | 0.541–0.870 | Moderate | 30 |
| Q6 | Standardization | 0.838 | 0.738–0.910 | Good | 30 |
| Q7 | Evidence Traceability | 0.869 | 0.787–0.928 | Good | 30 |
| Q8 | Innovation & Value | 0.901 | 0.835–0.947 | Excellent | 29<sup>a</sup> |
| Overall (pooled) |  | 0.843 | 0.812–0.870<sup>b</sup> | Good | 239 rows |

<sup>a</sup> One case lacks a Gemini Q8 value and is excluded from Q8 under the complete-case rule (Supplementary File S8). <sup>b</sup> Case-level cluster-bootstrap 95% interval: 0.782–0.874. Interpretation per Koo and Li [12]: $\geq 0.90$ excellent; 0.75–0.90 good; 0.50–0.75 moderate. 95% CI: absolute-agreement interval [18].

All materials—reports, scoring prompt, rubrics and defect taxonomy—are in Traditional Chinese and follow the ten-step QCC report convention of Taiwanese hospitals and the national healthcare quality award. Each report is a structured file of eight narrative fields (team, topic selection, current state, target setting, cause analysis, countermeasures, results, standardisation) of 323–1,317 Chinese characters (median 799); an English rendering of one case, a glossary and a crosswalk from dimensions to QCC steps and trap types are in Supplementary File S10.

The cases were drafted by Claude Opus 4.6 from a generation prompt that specified topic, requested quality level, traps and difficulty, and instructed the model to append the trap manifest and the eight anchor scores; the anchors are therefore the generator model’s own scores, constrained by the requested quality level. The first authorreviewed the first five cases in two rounds (ten edits) and spot-checked the remaining 25 without a written record; no anchor was independently re-derived by a human rater. Report length tracks the requested quality level (median 400, 812 and 1,185 characters for LOW, MEDIUM and HIGH; Pearson *r* with the mean anchor 0.878).

A comparator set of eight publicly available QCC reports from PubMed Central (PMC) was used for a descriptive plausibility check only (Supplementary Table S6). The evaluators did not receive the articles: each was rendered by the first author, with LLM assistance, from English reading notes into the same Traditional-Chinese template (705–825 characters), so the comparison is with condensed re-renderings, which also lack the “countermeasure *n* (addresses true cause *n*)” labelling of the synthetic template.

### Blinding of evaluator inputs

Evaluators received only the report text: the trap manifest and anchor scores are a trailing metadata block in each source file, every evaluator input was produced by truncating the source at that block, and the pipeline asserts before every run that no input contains a ground-truth marker. This check is reported explicitly because version 1 of this preprint did not meet it (see *Correction relative to version 1*).

### Evaluators and scoring

Five LLM evaluators were assessed: GPT (`openai/gpt-5.4`), Gemini (`google/gemini-3.1-pro-preview`), Grok (x-ai/grok-4. 6), DeepSeek (`deepseek/deepseek-r1`), and Claude (`anthropic/ claude-sonnet-4.5`). Per the pre-specified protocol the primary panel was *k*=4 (GPT, Gemini, Grok, DeepSeek); Claude was excluded because a same-family model generated the cases and was used in a pre-specified *k*=5 sensitivity analysis.

The scoring prompt (version 1.0, 1 May 2026) assigns the persona of an experienced Taiwanese healthcare-quality reviewer with national quality-award jury experience, defines the eight dimensions with five-level anchored rubrics, and requests a 1–5 score with quoted evidence per dimension plus a defect list drawn from a nine-category taxonomy with location and severity (severity was not analysed). Its user prompt lists nine methodological checks to perform before scoring, corresponding to nine of the ten trap types (all but Q06), so trap detection in this study is cued rather than unprompted. Two variants (v1.1, v1.2; 2–3 May 2026) drafted after the authors’ earlier pilot were tried informally with inter-model agreement as the criterion; neither was adopted and no outputs were archived.

All evaluators were called with temperature 0 and a 4,096-token completion limit through OpenRouter’s OpenAI-compatible API on 15 September 2026, three independent runs per evaluator per case; the median per case-dimension per evaluator is that evaluator’s summary score (run-to-run stability: Supplementary Table S3), and throttled requests were retried after the gateway’s stated delay. Some responses reached the completion limit before the score block or defect list was complete and contribute whatever was emitted before the cut (Supplementary Table S4, File S8); completion status and token usage were not stored (provenance: Supplementary File S7). Reporting follows TRIPOD-LLM [10] and GRRAS [11]; completed checklists are in Supplementary File S11.

### Statistical analysis

The primary endpoint was ICC(A,1)—intraclass correlation for absolute agreement, single measures—estimated across the four primary-panel evaluator medians for each dimension and for all case-dimension rows pooled [12, 13, 14, 15, 16, 17]; 95% confidence intervals use the absolute-agreement interval of McGraw and Wong [18], cross-checked against an independent implementation. ICCs ≥0.90 were classified as excellent, 0.75–0.90 as good, and 0.50–0.75 as moderate [12]. The protocol targeted a 95% CI half-width of about 0.15 for a dimension-level ICC of 0.75 with four evaluators (30–40 cases); observed half-widths were 0.06–0.16. Because the pooled analysis treats a case’s eight rows as independent, its analytic interval is anti-conservative, and a case-level cluster bootstrap (4,000 resamples of cases, percentile interval) is reported alongside it. The consensus score for a case-dimension is the median of the four primary-panel evaluator medians.

Secondary endpoints were (i) the *k*=5 sensitivity analysis adding Claude; (ii) leave-one-out sensitivity within the primary panel and a re-analysis under the score parser fixed at prompt development (see Missingness); (iii) anchor calibration—|consensus − anchor| ≤ 1 on trap-affected cells, with mean absolute error and exact-match rate because a *±*1 tolerance is lenient, and within each quality level and per evaluator (Supplementary Table S9); (iv) trap detection— whether an evaluator’s defect list contains a defect of the category expected for the planted trap (primary detector), with a keyword detector as a secondary signal and the trap-absent emission rate of the same category as a specificity check; and (v) a descriptive synthetic-versus-PMC check using Cliff’s delta [19] with a non-formal absolute-mean-difference margin of 1.0.

The trap-to-dimension map, expected defect category per trap type and keyword lists were fixed in a versioned file on 8 September 2026, before the scoring run, from the trap catalogue and trap descriptions only; because version control had been retired, the date rests on the file timestamp and project log, and the file is in the data release. Calibration is computed over the unique cells touched by at least one trap (58 traps on 52 cells). Detection is reported per run and per evaluator (at least one of three runs), with panel-majority and panel-any aggregations in the supplement because the aggregation level changes the estimate materially; defect lists were extracted with a parser that accepts the five list layouts the evaluators produced (Supplementary Methods).

### Missingness

A (case, dimension) row enters the pooled ICC only if all four primary-panel evaluators produced a median score for that cell; a run counts as parsed when its eight scores can be recovered from the response. The parser fixed at prompt development rejected 43 responses, 32 of them complete score blocks in layouts it had not anticipated; because these rejections followed evaluator layout rather than case content and removed three whole cases, we adopted a structure-aware parser as the primary analysis—a post hoc decision made after inspecting the responses—and report the original parser as a sensitivity analysis. Under the primary parser every case has a parsed run from every primary-panel evaluator; one case lacks a Gemini Q8 value, giving *n*=30 for Q1–Q7, 29 for Q8, and 239 pooled rows (details: Supplementary File S8).

## Results

### Inter-model reliability

Per-dimension ICC(A,1) on the primary *k*=4 panel ranged from 0.747 (Q5, Methodological Rigor) to 0.901 (Q8, Innovation and Value): six point estimates fell in the good range, Q5 just below it and Q8 in the excellent range per Koo and Li’s cutoffs [12], while the lower 95% confidence bounds of Q1, Q2, Q3, Q5 and Q6 extended into the moderate range (Table 1; Figure 1). The pooled ICC(A,1) across all case-dimension rows was 0.843 (95% CI 0.812 to 0.870; *n* = 239 rows, *k* = 4); the case-level cluster-bootstrap interval, which does not treat a case’s eight rows as independent, was 0.782 to 0.874. Across all 240 case-dimension cells the primary-panel consensus had Pearson *r* = 0.782 with the designed anchor, mean absolute error 0.48, exact agreement in 45.8% and agreement within *±*1 in 97.5%; 51.5% (1432/2779) of individual primary-panel run scores equalled the anchor exactly.

**Figure 1.**
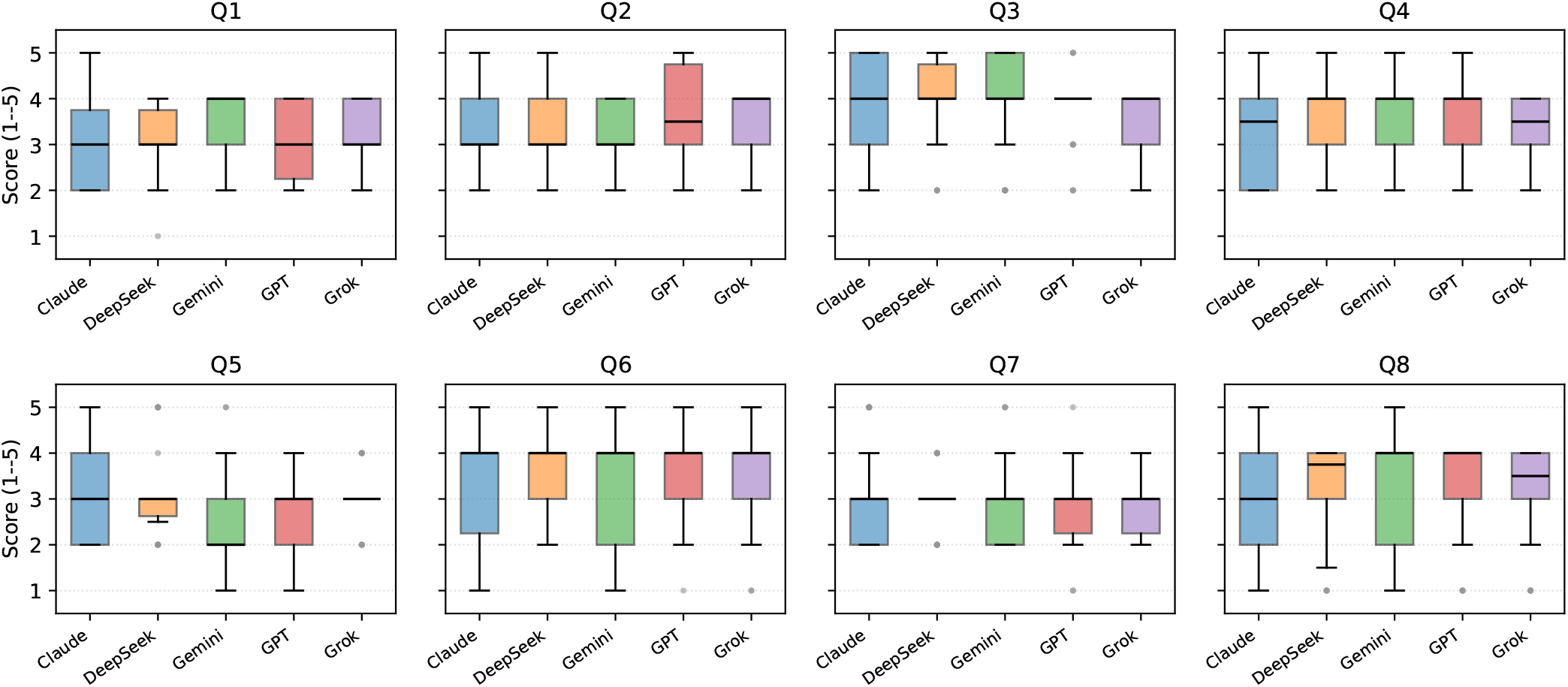
Per-dimension distribution of QCC quality scores across the 30 synthetic cases. Each box summarises one evaluator’s median-across-runs score on a given dimension; boxes are grouped by dimension (Q1–Q8) and coloured by evaluator. All five evaluators are shown to visualise the contribution of the sensitivity-only Claude evaluator alongside the four primary-panel evaluators (DeepSeek, Gemini, GPT, Grok); the primary ICC reported in Table 1 (0.843) uses only the primary panel; run-to-run stability per evaluator is in Supplementary Table S3. Box: 25th–75th percentile; horizontal line: median; whiskers: 1.5*×*IQR; grey points: out-of-whisker observations.

### Sensitivity analysis

The *k*=5 sensitivity adding Claude yielded a pooled ICC of 0.833 (Δ = −0.010 from the primary estimate), indicating that the same-family Claude evaluator did not inflate the pooled estimate. Leave-one-out estimates within the primary *k*=4 panel ranged from 0.835 (dropping GPT) to 0.857 (dropping Grok), confirming that no single evaluator drove the aggregate (Table 2). Re-analysis under the score parser fixed at prompt development, which excluded three cases, gave 0.854 (95% CI 0.824 to 0.881; *n* = 215 rows; Supplementary File S8).

**Table 2.** Sensitivity analyses around the primary *k*=4 endpoint (overall pooled case-dimension rows).

| Analysis | ICC(A,1) | Rows |
| --- | --- | --- |
| **Primary endpoint ($k=4$, Claude excluded)** | **0.843** | 239 |
| $k=5$ sensitivity (Claude included) | 0.833 | 239 |
| *Leave-one-out within primary panel ($k=3$):* |  |  |
| $k=3$, drop DeepSeek | 0.839 | 239 |
| $k=3$, drop Gemini | 0.839 | 240 |
| $k=3$, drop GPT | 0.835 | 239 |
| $k=3$, drop Grok | 0.857 | 239 |
| $k=4$, score parser fixed at prompt development | 0.854 | 215 |

### Anchor calibration and trap detection

Score-based anchor calibration was met in 50 of 52 trap-affected case-dimension cells (96.2%; Figure 2), exceeding the pre-specified 80% target. The two misses were the Q3 cell of QCC-P02 and the Q4 cell of QCC-P20 (which carries two traps), where the consensus exceeded the anchor by 2.0 and 1.5 points. Because a *±*1 tolerance is lenient on a five-point scale, the same 52 cells are also summarised directly: the consensus had a mean absolute error of 0.54 against the anchor, matched it exactly in 34.6% (18/52), and exceeded it by 0.24 points on average, both misses being overestimates. Both missed cells carry contestable anchors: in QCC-P02 the countermeasure adds a forced confirmation step in the hospital information system, which evaluators credited as a system-level action whereas the trap treats it as compliance enforcement. Calibration also varied with the requested quality level: across all cells the consensus ran 0.38 points above the anchor on LOW cases and 0.55 below on HIGH cases (exact agreement 36%, 59% and 31% for LOW, MEDIUM and HIGH), and agreement with the anchors was highest for the same-family Claude evaluator (exact 61%, mean absolute error 0.40) compared with 50–54% and 0.47–0.53 for the four primary-panel evaluators (Supplementary Table S9).

**Figure 2.**
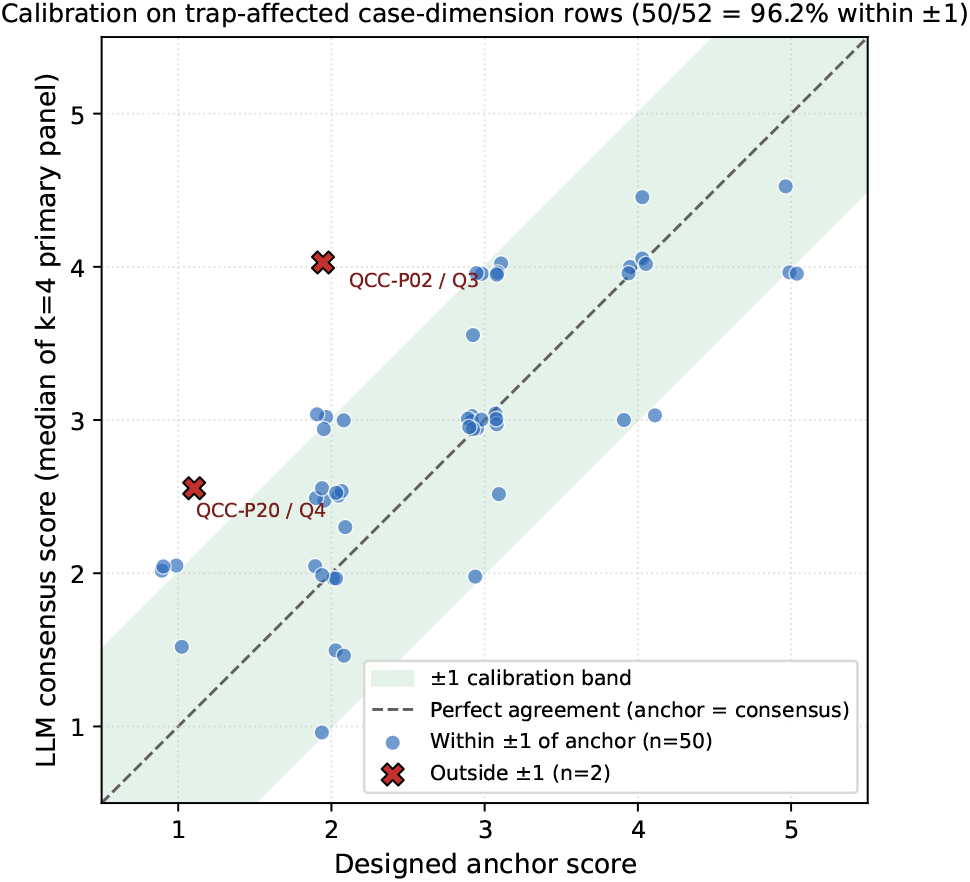
Designed-anchor vs LLM-consensus score on trap-affected case-dimension cells (*n* = 52 unique cells carrying 65 trap-dimension pairs). Each point is one cell; the LLM consensus is the median across the four primary-panel evaluator medians (GPT, Gemini, Grok, DeepSeek). Shaded band is the prespecified *±*1 calibration tolerance; the dashed line is perfect agreement. The two points outside the band are the Q3 cell of case QCC-P02 (+2.0) and the Q4 cell of case QCC-P20 (+1.5). Points are jittered slightly for visibility.

With the primary category detector, a defect of the expected category was present in 606 of 696 evaluator runs (87%; 92% of the 660 runs that returned any defect list) and was emitted by 217 of 232 (trap, evaluator) pairs (94%); the panel-majority rate was 98% (57/58) and the panel-any rate 100% (58/58). The keyword detector was more conservative at every level (55% per run; 68% per evaluator; 90% panel-any); requiring the defect’s location to match the planted section gave 64% per run and 77% per evaluator. Per-evaluator category detection by trap type ranged from 85% (Q07) to 100% (Q02, Q06, Q09); Q07 was detected by every evaluator in the two cases where standardisation was absent altogether (8/8 pairs) but missed three times in the three cases where a revised procedure was present and the planted weakness was its limited depth (9/12). The keyword detector was markedly weaker for Q04 (18%) and Q06 (25%), where evaluators named the defect in their own words. Specificity was limited: the expected category was also emitted for 64–98% of (case, evaluator) pairs in which the trap was absent, so the lift attributable to the planted trap was +2 to +6 percentage points for the four trap types that map to the generic methodology category and +14 to +21 points for the others (Supplementary Table S4).

### Synthetic-versus-PMC distributional plausibility

All eight dimensions met the pre-specified absolute-mean-difference margin of 1.0 (largest mean difference 0.49), but the two sets were not interchangeable (Table 3): synthetic consensus scores tended to be higher on Q1–Q5 (Cliff’s *d* = 0.125 to 0.475), with only the Q3 (Action Alignment) interval excluding zero (*d* = 0.475, 0.15 to 0.80) and the Q4 interval reaching it (0.00 to 0.67); Q6–Q8 showed negligible differences. Cross-set cosine similarity of the eight-dimension consensus profiles was 0.981 (within-synthetic 0.981; within-PMC 0.986).

**Table 3.** Synthetic-versus-PMC distributional plausibility check on the primary *k*=4 consensus (synthetic *n* = 30 cases, PMC *n* = 8; descriptive only, not an equivalence test).

| Code | Dimension | Synth $\bar{x}$ | PMC $\bar{x}$ | Cliff's $d$ (95% CI) | Effect | Within margin<sup>a</sup> |
| --- | --- | --- | --- | --- | --- | --- |
| Q1 | Root Cause Depth | 3.10 | 2.81 | 0.271 (−0.05, 0.57) | Small | Yes |
| Q2 | Systemic Coverage | 3.33 | 3.19 | 0.125 (−0.19, 0.43) | Negligible | Yes |
| Q3 | Action Alignment | 3.77 | 3.44 | 0.475 (0.15, 0.80) | Large | Yes |
| Q4 | Data Foundation | 3.49 | 3.00 | 0.346 (0.00, 0.67) | Medium | Yes |
| Q5 | Methodological Rigor | 2.81 | 2.50 | 0.225 (−0.15, 0.58) | Small | Yes |
| Q6 | Standardization | 3.48 | 3.62 | 0.075 (−0.25, 0.43) | Negligible | Yes |
| Q7 | Evidence Traceability | 2.95 | 2.94 | 0.000 (−0.33, 0.33) | Negligible | Yes |
| Q8 | Innovation & Value | 3.10 | 3.44 | −0.069 (−0.41, 0.32) | Negligible | Yes<sup>b</sup> |

<sup>a</sup> Margin check: $|\bar{x}_{\text{synth}} - \bar{x}_{\text{PMC}}| < 1.0$ ; descriptive plausibility check, not a formal equivalence test. <sup>b</sup> Synthetic $n = 29$ for Q8.
Cliff's $d$ [19] = $P(\text{synthetic} > \text{PMC}) - P(\text{synthetic} < \text{PMC})$ , so its sign can differ from that of the mean difference when ties are frequent (Q6); 95% CI by percentile bootstrap (4,000 resamples). Effect-size labels use the thresholds commonly applied to Cliff's $d$ ( $|d| < 0.147$ negligible; $< 0.33$ small; $< 0.474$ medium; otherwise large).

## Discussion

The main finding is good, but not near-perfect, inter-model reliability: four primary-panel LLM evaluators from different model families produced concordant QCC quality scores on a designed-anchor benchmark, with a pooled ICC(A,1) of 0.843 (95% CI 0.812–0.870) and per-dimension estimates between 0.75 and 0.90. The estimate was robust to the sensitivity analyses: adding the same-family Claude evaluator lowered it slightly (*k*=5 ICC = 0.833), leave-one-out within the primary panel ranged from 0.835 to 0.857, and the score parser fixed at prompt development, which excluded three cases, gave 0.854. These results are consistent with evidence that prompt consistency and structured scoring targets reduce LLM output variability [9], and place the achievable agreement in the good rather than the excellent band.

On trap-affected cells the consensus fell within *±*1 of the anchor in 96% of cells, but the mean absolute error was 0.54, only 35% of consensus scores matched the anchor exactly, and the dispersion was regressive—lenient on LOW cases, strict on HIGH cases—so evaluators scored in the intended direction without reproducing the designed values. Structured defect output was the stronger detection signal: the expected category appeared in 87% of evaluator runs and for 94% of (trap, evaluator) pairs, whereas the pre-specified keyword lists caught 55% and 68%, and as few as 18–25% for the two trap types that evaluators described in their own words. The category signal is not specific, however: the generic methodology category was emitted for almost every case whether or not a matching trap was present, so for the four trap types that map to it the detection rate is an upper bound, and requiring the defect’s location to match the planted section (77% per evaluator) is the more discriminating summary; future benchmarks should score detection on category and location fields and pair every rate with its trap-absent base rate. The Q07 pattern is instructive: evaluators always flagged a report with no standardisation at all but did not penalise a substantial standardisation section for omitting contingency procedures—a leniency human reviewers might share—so absence of content is under-reported only when partial.

These findings extend LLM-evaluation methodology to QCC quality assessment. Benchmarks in clinical knowledge [7], ophthalmology quality improvement [8] and chatbot health advice [20] compared a single LLM against a multiple-choice or human reference; the LLM-as-Judge paradigm [21] cautioned against single-judge self-preference and drift, which the multi-evaluator design and leave-one-out check partly address. Compared with QCC work that surveyed adoption [1] or quantified outcome effects [3, 2], this study targets the methodological scoring of the report itself.

The practical implication is a triage design to be validated, not a deployable tool. Multi-LLM scoring at ICC ≈ 0.84 could, if validated against expert judgment, augment expert QCC review by ranking submission batches, flagging reports below a defensible threshold for expert attention with the defect list attached, and supporting formative feedback in QCC training [22]; it cannot replace expert adjudication of individual reports. Such a design needs safeguards this study can only name: both calibration misses were overestimates, so the failure that matters is a weak report scored as adequate and not referred; case-level disagreement among evaluators should trigger human adjudication regardless of score; no report should be archived as exemplary on LLM scores alone; feedback should reach circles through their facilitators; a published rubric can be written to rather than met; and sending institutional reports, which name staff and units, to commercial APIs is a governance decision.

Output format was not stable between the authors’ earlier run (May 2026) and this one: DeepSeek responses accepted by the parser fixed at prompt development fell from 89 to 67 of 90 (inline comments on score lines; 41 of 90 responses in Simplified rather than Traditional Chinese) and Gemini’s from 81 to 70, although provider change, input priming and parser sensitivity cannot be separated because the earlier inputs were not blinded; and the Grok identifier used earlier (`x-ai/grok-4.1-fast`) was withdrawn by the gateway before this run. Multi-model panels inherit the release cadence of every member, so output format and model revision should be monitored per model over time; the leave-one-out stability (0.835–0.857) suggests a smaller panel may suffice for routine triage, and a formal evaluator-count optimisation under cost, latency and availability constraints is a natural next step.

The generalisability of these results has explicit limits. The anchors were drafted by the generator model and reviewed by one author, not derived by independent experts; they define a benchmark structure rather than a ground truth, and the same-family Claude evaluator agreed with them more closely than any primary-panel evaluator—a coupling on the anchor side that the *k*=5 analysis of inter-evaluator agreement cannot detect. Report length tracks the requested quality level (*r* = 0.88 with the mean anchor), so part of the calibration could be reproduced from length alone; a length-matched benchmark is needed to separate the two. Some traps encode conventions on which QCC practice is not uniform—a reversed but numerically equivalent achievement-rate formula, an undeclared alternative definition of improvement rate, a forced confirmation step scored as mere enforcement—so some “misses” are partly disagreements with the anchor. The benchmark is Traditional-Chinese and Taiwanese in format, persona and terminology; the traps cover five of eight dimensions, and one trap (fishbone categories omitted) is mapped to Q5 by the case specification although the rubric also penalises it under Q2; detection was cued by a prompt listing nine of the ten trap types; and LLM-generated cases may not capture the heterogeneity of real reports. Synthetic cases scored higher than the PMC comparators on five of eight dimensions, significantly so only for Action Alignment, but the comparators were condensed re-renderings without the countermeasure-to-cause labels of the synthetic template, so the difference partly reflects the rendering. Some responses were cut at the completion limit, completion status and token usage were not stored, and four responses received in an aborted first attempt were re-drawn (Supplementary File S7). Detection rates depend on the aggregation level (87% per run to 100% for any evaluator in any run), the keyword detector undercounts the evaluator that answered in Simplified Chinese, and the category detector’s high base rate makes the reported rates upper bounds. The study supports a reliability and a benchmark feasibility claim; it does not establish that LLM scores agree with expert judgment, reflect clinical utility, or generalise beyond the structured anchor context [23].

Reliability is a necessary precondition for valid assessment; the next step is an expert-labeled validation study on clinical QCC reports from practice, measuring accuracy against expert judgment alongside the consistency shown here and testing whether partial omissions, the pattern hardest to detect on the synthetic benchmark, are also under-reported in real-world reports.

## Conclusions

This designed-anchor pilot establishes that multi-model LLM scoring of QCC methodological quality achieves good inter-model reliability across eight quality dimensions, with a primary-panel (*k*=4) pooled ICC(A,1) of 0.843 and leave-one-out estimates between 0.835 and 0.857, and that evaluators identify planted methodological weaknesses in their structured output far more often than surface keyword matching detects. Benchmark feasibility is established; expert-labeled validation in clinical QCC reports is the planned next step.

### Correction relative to version 1

Version 1 of this preprint (posted 13 August 2026; doi: olin-kurl10.64898/2026.08.12.26360276v1) reported results from an evaluation run in which the evaluator inputs for 25 of the 30 synthetic cases still contained the trap manifest and the designed-anchor scores, so those cases were scored with the answer key inside the prompt. Version 1 should not be cited; every numerical result in its abstract, tables, figures and supplement is superseded by this version. An internal audit on 23 August 2026 identified the defect (evaluator scores matched the designed anchor exactly in 97% of dimension scores on the affected cases versus 40% on the five correctly blinded cases). Every number in version 1 that depended on the synthetic scores—the primary ICC of 0.953, the calibration, defect-mention and synthetic-versus-PMC results—was therefore invalid as reported. All 30 synthetic cases and the 8 PMC cases were re-scored on 15 September 2026 with blinded inputs verified programmatically. The same audit corrected further issues that also affected version 1: the trap-to-dimension map omitted the second dimension degraded by trap Q04; the calibration denominator counted trap instances rather than unique case-dimension cells; the keyword list used for defect mention was misassigned for five of ten trap types, which invalidates the version 1 interpretation of trap types Q05 and Q07; the ICC confidence intervals used a consistency-type formula rather than the absolute-agreement formula; per-evaluator medians of two runs were truncated rather than averaged; and the parser that extracted structured defect lists discarded or mislabelled a large share of items for two primary-panel evaluators. The score parser was also replaced, after inspection of its rejections, by one that accepts the score-block layouts the evaluators actually produced, so that all 30 cases enter the primary analysis; the original parser is retained as a sensitivity analysis. The headline detection metric therefore changed in definition as well as value: version 1 reported keyword mention pooled over all evaluators and runs (87.9%), whereas this version reports defect-category detection per run and per evaluator. Figures 1–2 and Supplementary Tables S1–S8 were regenerated, and the conclusion was revised from excellent to good inter-model reliability. The Grok evaluator identifier used in version 1 (`x-ai/grok-4.1-fast`) was withdrawn by the API gateway before the re-run and was replaced by `x-ai/grok-4.6`; the PMC comparator cases, scored once per evaluator in version 1, were scored three times to match the synthetic arm. This version also discloses matters that version 1 did not: the anchors were emitted by the generator model and reviewed by one author; the PMC comparators were scored as condensed Traditional-Chinese re-renderings, not as the published texts; the benchmark is Taiwanese in language and format; the prompt cues nine of the ten trap types; report length tracks the requested quality level; some responses were truncated at the completion limit; and the score parser was changed post hoc (Methods, Supplementary Files S8, S10 and S11). The version 1 outputs are retained in the project archive and are not used anywhere in this version.

## Data Availability

All data produced in the present study are available upon reasonable request to the authors

## Acknowledgments

The authors thank the quality-improvement community whose public materials make this kind of methodological work possible.

## Data Availability

This study used synthetic QCC reports and publicly available QCC reports from PubMed Central. No patient data were used. Data and materials supporting the findings are available from the corresponding author upon reasonable request. Public PMC source materials remain subject to the terms of their original publishers.

## Code Availability

The runner scripts, analysis scripts, figure-generation scripts, and the requirements.txt dependency pin file are available from the corresponding author upon reasonable request. Commercial LLM API access (OpenRouter) is required for an end-to-end re-run of the scoring pipeline; archived raw outputs supplied on request allow all manuscript and supplementary numbers to be regenerated without further API calls.

## Authors’ Contributions

HML conceived the study, developed the eight-dimension QCC quality framework, designed the synthetic-case trap manifest, developed and ran the multi-LLM scoring pipeline, conducted the statistical analyses, interpreted the results, and drafted the manuscript. JL provided methodological supervision, industrial- and information-management expertise, and critical review of the framework and manuscript. Both authors reviewed and approved the final manuscript.

## Use of AI in Manuscript Preparation

LLM-based scoring is the study’s subject of investigation, described in Methods. Separately, during manuscript preparation, the first authorused Anthropic Claude family models as a coding assistant and editing aid; all scientific claims and final wording were authored and approved by the human authors. AI tools are not listed as authors per ICMJE recommendations and applicable journal policy.

## Ethics Approval and Consent to Participate

Institutional review board approval and participant consent were not required because this methodological reliability study used LLM-generated synthetic QCC reports and publicly available QCC reports from PubMed Central. No identifiable patient-level institutional QCC reports were used.

## Consent for Publication

Not applicable.

## Abbreviations

ICC: intraclass correlation coefficient
LLM: large language model
LOO: leave-one-out
QCC: Quality Control Circle
PMC: PubMed Central
TRIPOD-LLM: Transparent Reporting of a multivariable prediction model for Individual Prognosis Or Diagnosis – Large Language Models

