## Supplementary for "Multi-model LLM assessment of Quality Control Circle methodological quality: a designed-anchor reliability study"

**Contents**

- SM1. Supplementary Methods (prompt and statistical detail)
- SM2. Supplementary Tables S1–S6 and S9
- SM3. Supplementary File S7 – Provenance Appendix
- SM4. Supplementary File S8 – Missingness and Parse Failures
- SM5. Supplementary File S10 – QCC report format, English rendering of one case, glossary and crosswalk
- SM6. Supplementary File S11 – TRIPOD-LLM and GRRAS reporting checklists

**Code, prompt, raw responses, and derived outputs:** available from the corresponding author upon reasonable request
**Suggested filename for submission:** `SupplementaryMaterial.pdf`

### SM1. Supplementary Methods

#### SM1.1. Prompt and scoring framework

The full evaluation prompt (`qcc-quality-eval-v1.md`, version 1.0, frozen 2026-05-01) is available from the corresponding author upon reasonable request. The prompt is written in Traditional Chinese. Its system part instructs the evaluator LLM to assume the role of a healthcare-quality expert with more than fifteen years' experience who has served on the jury of Taiwan's National Healthcare Quality Award and has QCC facilitation and root-cause-analysis review experience, to score eight QCC quality dimensions on a 1–5 ordinal anchor scale, to quote supporting text for each score, to treat any step not mentioned in the report as missing, and to produce a structured list of identified defects, each tagged with a category from a fixed nine-category taxonomy (DEF-ROOT, DEF-FACTOR, DEF-ACTION, DEF-DATA, DEF-METHOD, DEF-STANDARD, DEF-TIMELINE, DEF-LINK, DEF-INTANGIBLE), a severity (Critical/Major/Minor; requested but not analysed), a location and a description. Its user part lists nine checks to perform before scoring (candidate-topic comparison, target formula, objective cause verification, fishbone category coverage, improvement- and achievement-rate formulas, countermeasure-to-cause correspondence, comparability of pre/post observation periods, concreteness of standardisation, plausibility of intangible results); these correspond to trap types Q09, Q02, Q03, Q01, Q08, Q04, Q05, Q07 and Q10 respectively, so every trap type except Q06 (PDCA Check step missing) is cued by the prompt. Two later variants (v1.1, 2 May 2026, which added decision trees and a deduction scheme; v1.2, 3 May 2026, which reverted to the v1.0 anchors and added scoring examples) were drafted after the authors' earlier pilot run with inter-model agreement as the criterion; no outputs of those trials were archived, neither variant was adopted, and all runs reported here used v1.0 unchanged. The eight dimensions are Q1 root-cause depth, Q2 systemic factor coverage, Q3 action-cause alignment, Q4 data foundation, Q5 methodological rigour, Q6 standardisation, Q7 evidence traceability, and Q8 innovation and value. Each dimension carries five labelled anchor descriptions (Levels 1–5) defining what evidence in the report would warrant that score; these anchors are the same anchors against which calibration is measured in Section SM1.3.

#### SM1.2. Synthetic case generation

Synthetic QCC reports were produced by Claude Opus 4.6 using the generation prompt `qcc-generation-v1.md` against a structured case format spec (`qcc-case-format-spec.md`). The generation prompt specifies the topic category and hint, the requested quality level (HIGH, MEDIUM or LOW, with an indicative score band for each), the trap identifiers to plant and their difficulty (Easy/Medium/Hard), and ends with the instruction to output the case in the fixed YAML format "and append the trap manifest and the expected scores"; the eight anchor scores of every case are therefore the generator model's own scores for the case it had just written, constrained by the requested quality level. The first five cases were generated in an interactive session and reviewed by the first author in two rounds (ten edits, recorded in `PILOT-REVIEW.md`: unit corrections, two trap substitutions, one added trap, wording of trap descriptions); the remaining 25 were generated by an automated session against a batch specification that called for a spot-check of five to seven cases, which was done without a written record. No anchor score was independently re-derived by a human rater. No decoding temperature or seed was set or recorded, so generation is not reproducible bit-for-bit; the generated cases are the fixed artefact. Report length tracks the requested quality level: the blinded inputs contain a median of 400 (LOW), 812 (MEDIUM) and 1,185 (HIGH) Chinese characters, and the Pearson correlation between input length and the mean anchor across the 30 cases is 0.878 (between length and the mean primary-panel consensus, 0.844). Each synthetic case was instructed to (i) describe a realistic clinical-quality improvement topic, (ii) contain a fishbone diagram covering the five conventional categories (people, equipment, materials, methods, environment), (iii) include pre/post measurement targets, and (iv) optionally embed one or more designed traps from the trap manifest. Each case file carries a trailing metadata block (`trap_manifest:` and `expected_scores:`) recording which traps were planted, the report section each was planted in, and the designed anchor score for every dimension; this block is stripped from the evaluator input (main text, *Blinding of evaluator inputs*) and the dimension each trap type degrades is taken from the case-format specification. Claude Opus 4.6 generated the cases and Claude Sonnet 4.5 is one of the five evaluators; the primary panel therefore excludes Claude ($k$=4), and the pre-specified $k$=5 analysis adding Claude is the self-preference sensitivity (Table S1).

#### SM1.3. Designed-anchor protocol

Designed anchors are the expected scores attached to every case-dimension cell in the source file, emitted by the generator model as described above and fixed before scoring. For a case containing trap T affecting dimension D, the designed anchor is the score the evaluator is expected to produce on D if the trap is detected and scored according to the framework; anchors are a benchmark structure, not expert labels. The calibration metric is the fraction of unique trap-affected (case, dimension) cells for which $|\text{LLM consensus} - \text{designed anchor}| \leq 1$, with the prespecified target set at $\geq$ 80%. The $\pm$1 tolerance reflects the ordinal scale's resolution and is consistent with the within-1 agreement convention used in clinical rating studies; because it is lenient on a five-point scale, the mean absolute error and the exact-match rate ($|\Delta| = 0$) are reported alongside it in the main text.

#### SM1.4. Public PMC comparator set

The PMC comparator set ($n$ = 8) consists of public QCC reports retrieved from PubMed Central between 1 and 6 May 2026 using the search expression `("quality control circle" OR "QCC") AND (PDCA OR "plan-do-check-act") AND hospital`, filtered to full-text English-language reports describing a complete QCC cycle. Reports were screened by the lead author for fit against the framework's eight dimensions; reports describing partial cycles or non-QCC quality interventions were excluded. One additional retrieved report describing an integrated multi-method quality-management programme (PDCA combined with root-cause analysis and interdisciplinary team review) rather than a single QCC cycle was excluded on this criterion. The evaluators did not receive the article texts. For each article, structured English reading notes (problem, team, duration, quality tools, causes, interventions, results; 1,400–6,300 characters) were prepared on 1 May 2026 and rendered on 2 and 6 May 2026 into the eleven-field Traditional-Chinese template used for the synthetic cases (705–825 Chinese characters per case; the synthetic median is 799); both steps were carried out by the first author with the assistance of the same LLM coding assistant used for the pipeline (Claude). The renderings therefore condense each article, translate it, and impose the synthetic template's field structure; unlike the synthetic cases they do not carry the explicit "countermeasure $n$ (addresses true cause $n$)" labelling that the generation prompt imposes, one rendering carries the article's own limitations paragraph, and because the scoring prompt treats anything not mentioned as missing, condensation systematically disadvantages the comparators on dimensions that reward completeness. The synthetic-versus-PMC comparison in Table S5 is therefore a comparison between synthetic reports and condensed re-renderings of published reports, not between synthetic and published reports. The eight retained source articles are listed individually in Table S6. The set is too small for inferential comparison and is used only for descriptive plausibility (Table S5); it does not constitute an external validation set.

#### SM1.5. ICC(A,1) statistical model

We use the intraclass correlation coefficient under a two-way random-effects model with absolute agreement at the single-rater unit (ICC(A,1) in the McGraw–Wong convention; ICC(2,1) in the Shrout–Fleiss convention), implemented in the analysis scripts and cross-checked against `pingouin`'s `intraclass_corr`. Cases are random; evaluators are random; agreement (not consistency) is the target because the substantive claim is that two LLM evaluators produce similar absolute scores, not merely correlated rank orders. Pooled estimates collapse all eight dimensions into a single (case, dimension) row identifier (the *row* index) so that the primary ICC is computed on 239 complete-case rows $\times$ 4 evaluator columns. Per-dimension ICCs reported in Table S1 are estimated separately on each dimension's complete-case subset. 95% confidence intervals use the absolute-agreement interval of McGraw and Wong (1996, Table 7), which retains the between-evaluator mean square; the implementation was cross-checked against `pingouin.intraclass_corr` on every dimension (point estimates identical to four decimals, intervals identical to two). Because the pooled analysis treats the eight dimension rows of a case as independent, its analytic interval is anti-conservative; a case-level cluster bootstrap (4,000 resamples of cases carrying all their rows; percentile interval; `compute_k4_icc.py`) is reported alongside it: 0.782–0.874 for the $k$=4 primary estimate and 0.764–0.869 for $k$=5, against analytic intervals of 0.812–0.870 and 0.802–0.861.

#### SM1.6. Leave-one-out and $k$=5 sensitivity

The leave-one-out (LOO) sensitivity removes each of the four primary-panel evaluators in turn and recomputes the pooled ICC on the remaining three (Table S2); the LOO range expresses the largest change in the point estimate attributable to dropping any single evaluator. The $k$=5 analysis adding Claude (Table S1) is the pre-specified self-preference sensitivity, motivated by the family overlap between the Claude Opus generator and the Claude Sonnet evaluator; adding Claude lowered rather than raised the pooled ICC (0.843 with $k$=4 vs 0.833 with $k$=5), so the inter-evaluator agreement claim does not rely on within-family coupling (agreement with the anchors themselves is a separate question; see main text).

#### SM1.7. Trap detection: category and keyword detectors

The trap detection map (`trap-keyword-map-v2.yaml`, fixed 8 September 2026, before any scoring run reported here) records for each of the ten trap types the report section in which it is planted, the quality dimension(s) it degrades (from the case-format specification), the defect category an evaluator is expected to emit, and a list of keyword patterns. The map was built only from the trap catalogue and the ground-truth trap descriptions in the case files; evaluator output was not consulted, so the map cannot have been tuned to the results it scores. Against those ground-truth descriptions every keyword list has 100% sensitivity for its own trap type and no other type's list reaches that level (validation script archived with the map).

*Category detector (primary).* The scoring prompt requires each defect in `defects_identified` to carry a `type` drawn from a fixed taxonomy (DEF-ROOT, DEF-FACTOR, DEF-ACTION, DEF-DATA, DEF-METHOD, DEF-STANDARD, DEF-INTANGIBLE, . . . ). A trap is detected in a given run if that run emits a defect whose type is in the trap's expected set. Eight trap types map to a single category; Q08 accepts two because the prompt taxonomy lists "calculation error" under both DEF-DATA and DEF-STANDARD, and Q09 accepts two because a topic chosen without data support can be reported either as a data problem (DEF-DATA) or as a skipped methodological step (DEF-METHOD). A stricter variant additionally requires the emitted defect's `location` to match the planted section.

*Keyword detector (secondary).* A trap is detected if any of its keyword patterns appears in the concatenated type, location and description text of the run's defects. Because the patterns come from the trap descriptions rather than from evaluator language, this detector is a conservative lower bound on mention.

*Aggregation.* Both detectors are reported at four levels: per run; per evaluator (detected in at least one of that evaluator's three runs; the headline level in the main text); panel majority (at least three of the four primary evaluators); and panel-any (at least one evaluator in at least one run, the union rule used in version 1 of this preprint). Runs whose score block failed to parse still contribute their defect list when a response was received; runs whose response was truncated before the defect list (18 Gemini, 4 DeepSeek, 11 Claude runs) count as non-detections at the run level and are also reported excluded. Defect lists were extracted from the archived responses with a structure-aware parser (`parse_defects_v2`) that locates the defect list under whatever key the evaluator used, splits it on list-item markers, and reads every DEF-* token from the item's category-bearing keys (`type`, `code`, `defect_code`, …, including list syntax); the original parser recognised only three key names and kept one key per item, which left 30 of 90 Grok runs with an empty list and mislabelled most GPT items. Keyword patterns are Traditional Chinese; DeepSeek answered in Simplified Chinese in 41 of 90 runs, which depresses its keyword rate but not its category rate.

#### SM2. Supplementary Tables

Table S1: Per-dimension ICC(A,1) on the $k$=5 sensitivity panel (full panel including Claude). The primary $k$=4 per-dimension ICCs are reported in main-text Table 1.

| # | Dimension | ICC(A,1) | 95% CI | $n$ | $k$ |
|---|---|---|---|---|---|
| Q1 | Root Cause Depth | 0.808 | [0.695, 0.892] | 30 | 5 |
| Q2 | Systemic Coverage | 0.789 | [0.679, 0.879] | 30 | 5 |
| Q3 | Action Alignment | 0.814 | [0.705, 0.896] | 30 | 5 |
| Q4 | Data Foundation | 0.846 | [0.759, 0.913] | 30 | 5 |
| Q5 | Methodological Rigor | 0.746 | [0.563, 0.865] | 30 | 5 |
| Q6 | Standardization | 0.834 | [0.741, 0.906] | 30 | 5 |
| Q7 | Evidence Traceability | 0.841 | [0.751, 0.910] | 30 | 5 |
| Q8 | Innovation & Value | 0.884 | [0.813, 0.936] | 29 | 5 |
| **Overall (pooled), $k$=5 sensitivity** | | **0.833** | **[0.802, 0.861]** | 239 | 5 |
| Overall (pooled), $k$=4 primary (cf. Table 1) | | 0.843 | [0.812, 0.870] | 239 | 4 |

ICC(A,1) = two-way random, absolute agreement, single measures. $n$ = complete-case count. $k$ = number of evaluators. 95% CI: McGraw–Wong absolute-agreement interval. Adding the same-family Claude evaluator lowers the pooled estimate by 0.010, indicating no self-preference inflation of the primary endpoint. Case-level cluster-bootstrap 95% intervals for the pooled estimates: 0.782–0.874 ($k$=4) and 0.764–0.869 ($k$=5).

Table S2: Leave-one-out ICC sensitivity within the primary $k$=4 panel (drop one of the four primary evaluators in turn; remaining $k$=3 pooled estimate).

| Dropped Evaluator | Remaining $k$ | Overall ICC(A,1) | Rows |
|---|---|---|---|
| DeepSeek | 3 | 0.839 | 239 |
| Gemini | 3 | 0.839 | 240 |
| GPT | 3 | 0.835 | 239 |
| Grok | 3 | 0.857 | 239 |
| **Full primary panel** | **4** | **0.843** | 239 |

Leave-one-out ICC range within the primary panel: 0.835–0.857 ($\Delta$ from primary: −0.008 to +0.014). Dropping Gemini restores the one case-dimension row (QCC-P30, Q8) that lacks a Gemini value. The $k$=5 sensitivity adding the same-family Claude evaluator yields ICC = 0.833; the score parser fixed at prompt development yields 0.854 on 215 rows (File S8).

Table S3: Intra-Model Stability Across 3 Runs

| Model | Mean Range | Max Range | Exact Agreement % | Comparisons |
|---|---|---|---|---|
| Claude | 0.23 | 2 | 78.3% | 240 |
| DeepSeek | 0.39 | 2 | 62.5% | 240 |
| Gemini | 0.13 | 2 | 87.3% | 212 |
| GPT | 0.18 | 1 | 81.7% | 240 |
| Grok | 0.13 | 1 | 86.7% | 240 |

Range = max − min score across the parsed runs for each case-dimension. Exact agreement = percentage of case-dimension pairs with identical scores across all parsed runs. Comparisons = number of case-dimension pairs with at least two parsed runs (30 cases × 8 dimensions, minus parse failures). Temperature was 0 for all evaluators; run-to-run variation therefore reflects provider-side non-determinism.

Table S4: Trap detection by trap type and aggregation level, primary $k$=4 panel (map v2, defect parser v2)

| | | | | Category detector | | | | Keyword detector | |
|---|---|---|---|---|---|---|---|---|---|
| Trap | Type | Freq | Dim | Evaluator | Majority | Any | Absent† | Evaluator | Any |
| Q01 | Fishbone incomplete | 6 | Q5 | 92% | 6/6 | 6/6 | 74% | 50% | 6/6 |
| Q02 | Goal without baseline | 6 | Q5 | 100% | 6/6 | 6/6 | 94% | 96% | 6/6 |
| Q03 | Cause verification skipped | 8 | Q5 | 97% | 8/8 | 8/8 | 94% | 94% | 8/8 |
| Q04 | Misaligned countermeasures | 7 | Q3,Q5 | 89% | 7/7 | 7/7 | 74% | 18% | 3/7 |
| Q05 | Non-comparable pre/post | 6 | Q4 | 88% | 5/6 | 6/6 | 81% | 71% | 5/6 |
| Q06 | Missing Check step | 5 | Q5 | 100% | 5/5 | 5/5 | 94% | 25% | 4/5 |
| Q07 | SOP not established | 5 | Q6 | 85% | 5/5 | 5/5 | 64% | 75% | 5/5 |
| Q08 | Achievement formula error | 6 | Q6 | 96% | 6/6 | 6/6 | 82% | 71% | 6/6 |
| Q09 | Topic without data support | 5 | Q4 | 100% | 5/5 | 5/5 | 98% | 100% | 5/5 |
| Q10 | Intangible benefits inflated | 4 | Q8 | 88% | 4/4 | 4/4 | 73% | 88% | 4/4 |
| **All 58 trap instances** | | | | **94%** | 98% | 100% | – | 68% | 90% |
| counts | | | | 217/232 | 57/58 | 58/58 | | 158/232 | 52/58 |
| Per run (696 evaluator-runs) | | | | 87% | – | – | | 55% | – |
| Per run, excluding 36 runs with no defect list | | | | 92% | – | – | | | |
| Category + planted location, per evaluator | | | | 77% | 79% | 90% | | | |

Freq = number of cases containing the trap. Dim = dimension(s) degraded per the case-format specification (trap Q04 degrades two). Evaluator = share of (trap instance, evaluator) pairs detected in at least one of that evaluator's three runs. Majority = trap instances detected by at least three of the four primary evaluators. Any = detected by at least one evaluator in at least one run (the union rule used in version 1). †Absent = share of (case, evaluator) pairs in which the same category was emitted although the case does *not* contain the trap (specificity check); the difference between the Evaluator and Absent columns is the lift attributable to the planted trap. Score-based calibration ($|$consensus − anchor$| \leq 1$) was met on 50 of the 52 unique trap-affected cells; the two misses were QCC-P02/Q3 (trap Q04, +2.0) and QCC-P20/Q4 (traps Q05+Q09, +1.5). Trap Q07 comprises two kinds of instance: in QCC-P07 and QCC-P12 no standardisation was established at all (8/8 evaluator pairs detected), whereas in QCC-P16, QCC-P21 and QCC-P26 a revised procedure is present and the planted weakness is its limited depth (contingency and cross-unit procedures omitted; 9/12 pairs, the three misses being Gemini on P16 and P21, two of whose runs were truncated, and DeepSeek on P26).

Table S5: Synthetic vs PMC Distributional Plausibility (primary $k$=4 consensus)

| Dim | Synth Mean | PMC Mean | Cliff's $\delta$ (95% CI) | Effect Size | Margin |
|---|---|---|---|---|---|
| Q1 | 3.10 | 2.81 | 0.271 (−0.05, 0.57) | Small | PASS |
| Q2 | 3.33 | 3.19 | 0.125 (−0.19, 0.43) | Negligible | PASS |
| Q3 | 3.77 | 3.44 | 0.475 (0.15, 0.80) | Large | PASS |
| Q4 | 3.49 | 3.00 | 0.346 (0.00, 0.67) | Medium | PASS |
| Q5 | 2.81 | 2.50 | 0.225 (−0.15, 0.58) | Small | PASS |
| Q6 | 3.48 | 3.62 | 0.075 (−0.25, 0.43) | Negligible | PASS |
| Q7 | 2.95 | 2.94 | 0.000 (−0.33, 0.33) | Negligible | PASS |
| Q8 | 3.10 | 3.44 | −0.069 (−0.41, 0.32) | Negligible | PASS |

Synthetic $n = 30$ cases (29 for Q8); PMC $n = 8$. Cliff's $\delta$ thresholds: $|\delta| < 0.147$ negligible, 0.147–0.33 small, 0.33–0.474 medium, $> 0.474$ large; positive values mean synthetic scores are higher; 95% CI by percentile bootstrap (4,000 resamples). Margin check: descriptive |mean difference| $< 1.0$ (not a formal TOST equivalence test). Cosine similarity of eight-dimension consensus profiles: within-synthetic 0.981, within-PMC 0.986, cross 0.981. All 8 dimensions passed the margin check, but synthetic scores exceeded PMC scores on Q1–Q5; only the Q3 interval excludes zero.

Table S6: Public PMC comparator set ($n = 8$): source articles scored in the descriptive plausibility check (Table S5).

| PMC ID | QCC topic | Source |
|---|---|---|
| PMC7249848 | Laboratory blood-specimen quality | *Medicine* 2020;99(21):e20333 |
| PMC7509727 | Biofilm formation in flexible endoscopes | *J Int Med Res* 2020;48(9):300060520952983 |
| PMC8519257 | Functional exercise execution after orthopaedic surgery | *Medicine* 2021;100(41):e27514 |
| PMC10783305 | Postoperative nursing after dental implant surgery | *Medicine* 2024;103(2):e36894 |
| PMC10994446 | Early ambulation after caesarean section | *Medicine* 2024;103(14):e37633 |
| PMC11523946 | Specimen rejection rate | *J Multidiscip Healthc* 2024;17:4925–4935 |
| PMC12046524 | Foreign objects retained in sterile packages | *Risk Manag Healthc Policy* 2025;18:1441–1454 |
| PMC12155372 | Surgical instrument pre-treatment failure | *Risk Manag Healthc Policy* 2025;18:1837–1845 |

All eight articles were openly available in PubMed Central before this study began; no request, application, screening, or registration was required for access. Full text for each is reachable at `https://pmc.ncbi.nlm.nih.gov/articles/` followed by the PMC ID (for example, `https://pmc.ncbi.nlm.nih.gov/articles/PMC7249848/`). In the deposited evaluation inputs these cases carry identifiers of the form `REAL-PMC7249848`. Bibliographic details were verified against the corresponding NCBI records. These articles are analysed source material rather than cited literature and are therefore listed here instead of in the manuscript reference list; reuse of the source articles remains subject to the terms of their original publishers.

Table S9: Agreement with the designed anchors by evaluator and by requested quality level (all 240 case-dimension cells; per-evaluator median across runs; primary-panel consensus for the quality-level rows)

| | Cells | Exact | Within ±1 | MAE | Signed error | $r$ |
|---|---|---|---|---|---|---|
| *By evaluator (median of three runs vs anchor)* | | | | | | |
| GPT | 240 | 52.5% | 97.1% | 0.50 | +0.10 | 0.751 |
| Gemini | 239 | 49.8% | 97.1% | 0.53 | +0.11 | 0.760 |
| Grok | 240 | 50.0% | 97.1% | 0.53 | +0.07 | 0.720 |
| DeepSeek | 240 | 53.8% | 97.9% | 0.47 | +0.20 | 0.788 |
| Claude[a] | 240 | 61.3% | 98.3% | 0.40 | +0.15 | 0.835 |
| *By requested quality level (primary $k$=4 consensus vs anchor)* | | | | | | |
| LOW (10 cases) | 80 | 36.2% | – | 0.57 | +0.38 | – |
| MEDIUM (14 cases) | 112 | 58.9% | – | 0.37 | +0.19 | – |
| HIGH (6 cases) | 48 | 31.2% | – | 0.59 | −0.55 | – |

[a]Sensitivity evaluator; same model family as the generator that emitted the anchors. Signed error = evaluator (or consensus) minus anchor; positive values mean scores above the anchor. The Claude evaluator, which shares a model family with the generator that emitted the anchors, agrees with the anchors more closely on every measure than any primary-panel evaluator; the $k$=5 sensitivity analysis (Table S1) tests inter-evaluator agreement and cannot detect this anchor-side coupling. Across quality levels the consensus is lenient on LOW cases and strict on HIGH cases, i.e. regressive toward the middle of the scale; input length is strongly associated with the requested level (Supplementary Methods), so part of this pattern may be reproducible from length alone.

### SM3. Supplementary File S7 – Provenance Appendix

#### SM3.1. Study Metadata

| Item | Value |
|---|---|
| Study | Q01 / Sub-study 1a |
| Design | Designed-anchor reliability benchmark |
| Synthetic cases | 30 |
| Public PMC comparator cases | 8 |
| Evaluators | GPT, Gemini, Grok, DeepSeek, Claude |
| Runs per evaluator | 3 (synthetic and PMC) |
| Scoring window | 2026-09-15 (corrected run; version 1 run was 2026-05-06/10) |
| Route | OpenRouter OpenAI-compatible chat completions |
| Prompt | `qcc-quality-eval-v1.md` (v1.0, 2026-05-01) |

#### SM3.2. Model Provenance

| Evaluator | Provider Model ID | Route | Role |
|---|---|---|---|
| GPT | `openai/gpt-5.4` | OpenRouter | Primary |
| Gemini | `google/gemini-3.1-pro-preview` | OpenRouter | Primary |
| Grok | `x-ai/grok-4.6`† | OpenRouter | Primary |
| DeepSeek | `deepseek/deepseek-r1` | OpenRouter | Primary |
| Claude | `anthropic/claude-sonnet-4.5` | OpenRouter | $k$=5 sensitivity |

†Version 1 used `x-ai/grok-4.1-fast`, which the gateway withdrew before the corrected run.

Generator model (synthetic case creation): Claude Opus 4.6 (`anthropic/claude-opus-4-6`). The generator shares a model family with the Claude Sonnet evaluator; the primary panel excludes Claude and the $k$=5 analysis adding it is the pre-specified self-preference sensitivity.

#### SM3.3. Run Configuration

Temperature = 0. Max completion tokens = 4096. Three independent runs per evaluator per case (synthetic and PMC); exact median aggregation across parsed runs before estimating ICC. Requests refused by the gateway with a throttling status (HTTP 402 in-flight budget, 429) were retried after the gateway's stated delay, up to 15 attempts; every reported call eventually succeeded or failed on content, never on throttling. Each case result was written to disk immediately on receipt. Total gateway usage for the corrected run, including one aborted attempt, was USD 46.77.

#### SM3.4. Software Environment

Python 3.13.9. The exact third-party dependencies actually imported by the analysis and figure scripts are listed in `04-analysis/sub-study-1/phase1-notebooks/requirements.txt` in the data release: numpy 2.4.0, scipy 1.17.1, matplotlib 3.10, PyYAML 6.0, python-dotenv 1.1.0, openai 2.32.0, and requests 2.32.5. Pinned with compatible-release operators (`~=`) to block minor-version bumps that could shift ICC point estimates.

#### SM3.5. Repository Snapshot

The project retired git-based version control on 23 August 2026; the last commit before the corrected run, `f7ce7cf`, is archived in the private repository `https://github.com/R78101029/study_QI_LLM` and contains the version 1 pipeline in the state that was audited. The corrected pipeline, the blinded evaluator inputs, the frozen trap map, the raw model outputs of the corrected run, the derived score matrices under both parsers, and the analysis scripts are archived as a versioned package (`q01_v2`) available from the corresponding author upon reasonable request; its `MANIFEST.json` lists every file with its SHA-256 hash, so the exact inputs to every number in this version can be verified without a version-control system. The version 1 outputs are retained in the project archive under a directory marked as contaminated and are not used anywhere in this version.

#### SM3.6. Provenance Gaps and Reproducibility Caveat

Provider-side model release snapshots, API request timestamps, and deployment revision hashes were not captured in this run; nor were the gateway's per-response `model`, `provider`, `id`, `finish_reason` or `usage` fields, so a response cut at the 4,096-token completion limit cannot be distinguished from other early terminations except by inspection (Gemini responses that ended before any complete score list are 259–1,564 characters long), and the gateway was free to route each request to any provider hosting the model identifier. The first attempt of the corrected run (six parallel processes) was aborted when the gateway refused requests for exceeding the in-flight credit budget; responses already persisted were reused in the second attempt for GPT (43 runs) and Gemini (16 runs) while the other three evaluators were re-run in full, and because resumption was keyed on a parseable score block, four responses received in the first attempt that had content but no parseable score block (DeepSeek QCC-P02 run 1; Gemini QCC-P16, P19 and P26 run 1) were re-drawn, so those four cells had a second chance that no other cell had. Re-running the OpenRouter pipeline against the same routing IDs at a later date may therefore return different responses if any provider has silently rolled forward to a successor checkpoint. Reproducibility is best described as *artifact-level*: the raw model outputs, parse rules, prompts, designed-anchor scores, and analysis code are archived in the data release, so all numbers in the manuscript and supplementary can be regenerated bit-identical from those archived inputs, but a fresh end-to-end re-run starting from API calls is not guaranteed to land on the same checkpoints. Between the May 2026 and September 2026 runs the share of DeepSeek score blocks accepted by the parser fixed at prompt development fell from 89/90 to 67/90 and Gemini's from 81/90 to 70/90, while GPT's were accepted in every synthetic run in both windows (Grok is a different model in the two windows); see File S8 for what changed in the responses and why the counts cannot separate provider change from parser sensitivity. Local artifact capture times are documented in `q01-provenance-appendix.md` in the data release.

### SM4. Supplementary File S8 – Missingness and Parse Failures

#### Complete-case construction algorithm

For each (case, evaluator), the per-evaluator median is computed across the (up to) three independent runs available for that pair, conditional on at least one run producing a parseable score for the target dimension. A (case, dimension) row enters the primary-panel pooled ICC denominator only if all four primary-panel evaluators (GPT, Gemini, Grok, DeepSeek) produced a usable median value for that cell; otherwise the row is excluded from that dimension's ICC. This is a complete-case rule applied at the row level after run aggregation, not a global case-level exclusion: a case that yields complete Q1–Q6 cells but missing Q7 contributes to the Q1–Q6 ICCs but not to Q7.

#### Two score parsers

The parser fixed at prompt development (`parse_scores`) is a cascade of line-anchored regular expressions written against the score-block layouts observed while the prompt was developed; it is not a schema validator. When applied to the corrected run it rejected 23 DeepSeek and 20 Gemini responses out of 90 each. Inspection showed that 22 of the DeepSeek rejections were complete answers under a `dimension_scores:` key—the same key that 57 of its May runs used and that the parser accepted—whose score lines now carried an inline comment (`Q1: 4 # ...`) that the line-anchored pattern does not accept, and that 10 of the Gemini rejections were responses that ended before the closing code fence but contained a complete score list (half of them under an `evaluations:` key). Because these rejections depended on the evaluator's layout rather than on case content, and because they removed three whole cases from a 30-case design, a structure-aware parser (`parse_scores_v2`) that accepts these layouts was adopted as the primary analysis on 17 September 2026, after the responses had been inspected; it is therefore a post hoc analysis decision, and the original parser is retained as a sensitivity analysis. Both parsers are applied to the same archived responses (`reparse_scores.py`); every per-run record carries both results.

#### Per-evaluator parse counts (synthetic 30-case run, 2026-09-15)

| | | Primary parser | | | Parser fixed at prompt development | | |
|---|---|---|---|---|---|---|---|
| Evaluator | Runs | Parsed | Partial | Unparseable | Parsed | Partial | Unparseable |
| Claude | 90 | 90 | 1 | 0 | 90 | 1 | 0 |
| DeepSeek | 90 | 89 | 2 | 1 | 67 | 2 | 23 |
| Gemini | 90 | 80 | 6 | 10 | 70 | 3 | 20 |
| GPT | 90 | 90 | 0 | 0 | 90 | 0 | 0 |
| Grok | 90 | 90 | 0 | 0 | 90 | 0 | 0 |

"Parsed" = at least one dimension score recovered; "Partial" = parsed but missing one or more of the eight dimension scores (Gemini's partial runs are responses truncated inside the score list); "Unparseable" = no dimension value recovered. Under the primary parser the single DeepSeek failure (QCC-P10 run 2) is an empty completion and the ten Gemini failures are responses truncated before any complete score list; these are genuine output failures rather than layout mismatches. For the PMC comparator cases (8 cases × 3 runs), parsed runs under the primary parser were Claude 24, GPT 24, Grok 24, Gemini 22, DeepSeek 24, and every PMC case had at least one parsed run from every evaluator; under the parser fixed at prompt development they were 24, 24, 23, 18 and 18, and one PMC case had no parsed Gemini run.

#### Sensitivity analysis under the parser fixed at prompt development

Under the original parser three cases (QCC-P10 from DeepSeek; QCC-P16 and QCC-P24 from Gemini) had no parsed run from one primary-panel evaluator and were excluded from every dimension, giving $n$=27 for Q1–Q7, 26 for Q8 and 215 pooled rows. The pooled $k$=4 ICC(A,1) was 0.854 (95% CI 0.824–0.881; case-level cluster bootstrap 0.790–0.882), $k$=5 0.842 (0.811–0.870), leave-one-out 0.844–0.863, and the per-dimension estimates were Q1 0.821, Q2 0.813, Q3 0.819, Q4 0.857, Q5 0.777, Q6 0.848, Q7 0.864 and Q8 0.917, each within 0.03 of the primary estimate (`compute_k4_icc.py` on `qcc-score-matrix-strict.json`). Under that parser the PMC comparison used seven PMC cases and 27 synthetic cases; all eight dimensions still met the margin and only Q3 excluded zero. The headline conclusion does not depend on the choice of parser; the primary parser was adopted so that no case is excluded because of an evaluator's layout.

#### Consequence on the pooled ICC denominator

Under the primary parser every case has at least one parsed run from every primary-panel evaluator; one case (QCC-P30) lacks a Gemini Q8 value because all three Gemini runs were truncated before that line. Q1–Q7 therefore retain $n = 30$ rows and Q8 $n = 29$, and the pooled $k$=4 primary-panel denominator is $30 \times 7 + 29 = 239$ rows (matching Table S1 and Table S2). The remaining missingness is a single row; dropping Gemini restores it ($n = 240$, ICC 0.839) without changing the estimate materially.

### SM5. Supplementary File S10 – QCC report format, English rendering of one case, glossary and crosswalk

#### Report format

Every synthetic case and every PMC rendering is a YAML file with three identifiers (`case_id`, `topic_category`, `circle_name`) and eight narrative fields that follow the order of the ten-step QCC cycle used in Taiwanese hospital quality-award reports: `team_composition`, `topic_selection`, `current_status`, `target_setting`, `root_cause_analysis`, `countermeasures`, `results`, `standardization`. The activity plan and the implementation step of the ten-step cycle are folded into the countermeasures field, and the review step into the standardisation field. Source files additionally carry the trailing `trap_manifest` and `expected_scores` blocks that are stripped from evaluator inputs.

#### English rendering of case QCC-P01 (blinded evaluator input)

The rendering below is a faithful translation by the authors of the evaluator input for QCC-P01 (MEDIUM quality; two planted traps), followed by the withheld trap manifest. Numbers, formulas and section order are unchanged.

**Circle:** "Clean Hands Circle"; topic category: infection control.

**Team composition.** Seven members: one infection-control nurse (circle leader), one medical-ward head nurse (facilitator), two N3 nurses (deputy leader and secretary), two N2 nurses, one environmental-cleaning team leader. General medical ward (40 beds), day-shift nurse-to-patient ratio about 1:8.

**Topic selection.** In response to the hospital's annual patient-safety goal "implement infection-control measures", the quality-management centre recommended that units run hand-hygiene improvement projects. The members voted for this topic; scores: hospital policy 5, importance 4, urgency 4, circle capability 4 (total 17/20). Infection-control committee statistics showed the ward's hand-hygiene audit compliance for the first half of 2024 was only 62%, below the hospital average of 75%.

**Current status.** Survey period 1 July–31 August 2024 (8 weeks). The infection-control nurse and the secretary observed 360 hand-hygiene opportunities by random-time observation. Overall non-compliance 37.8% (136/360). Pareto analysis: (1) before patient contact, 52 (38.2%, cumulative 38.2%); (2) after contact with patient surroundings, 35 (25.7%, 63.9%); (3) before aseptic/clean procedures, 24 (17.6%, 81.6%); (4) after patient contact, 15 (11.0%, 92.6%); (5) after body-fluid exposure, 10 (7.4%, 100%). The first three items, cumulative 81.6%, are the improvement focus.

**Target setting.** Target = current value − (current value × improvement focus × circle capability) = 37.8% − (37.8% × 81.6% × 0.72) = 37.8% − 22.2% = 15.6%. After discussion the members set the non-compliance target at 16% (slightly conservative).

**Root cause analysis.** Fishbone analysis in five categories. [Man] (a) nurses tend to skip hand-hygiene moments at busy times (medication and dressing rounds); (b) new N1 nurses are unfamiliar with recognising the "five moments". [Machine] (a) only two sinks in the ward corridor, off the main care route; (b) some wall-mounted alcohol-rub holders not refilled during maintenance. [Material] (a) after the hospital switched alcohol-rub brand, nurses reported a pungent smell and dry hands. [Method] (a) audit results published only monthly, feedback delayed 18 days on average; (b) no immediate positive-feedback mechanism. [Environment] (a) patient rooms are cramped; medication carts cannot reach the corridor sinks easily. True-cause verification: two weeks of checklist observation confirmed three true causes: (1) a knowledge–behaviour gap for the before-patient-contact moment (80 checks, only 48% compliant); (2) insufficient access to alcohol rub (unreachable within three steps at 60% of care-route locations); (3) delayed audit feedback weakens behaviour correction (interviews with 10 nurses, 8 of whom no longer remembered the situation when the report arrived).

**Countermeasures.** Countermeasure 1 (addresses true cause 1): produce a "five moments" pocket card for all nursing staff and run a five-minute scenario quiz at each weekly morning meeting. Countermeasure 2 (addresses true cause 2): add a clip-on alcohol-rub holder at every bed (fixed to the IV pole or bed rail) so that alcohol rub is within two steps at 90% of care-route locations. Countermeasure 3 (addresses true cause 3): introduce immediate feedback—the infection-control nurse posts audit results (anonymised) to the unit's LINE group on the day of the audit, and a weekly compliance trend chart is posted at the nursing station. Evaluation matrix (feasibility/effect/economy, 1–5 each): countermeasure 1, 11 points; 2, 13; 3, 12. All adopted. Implementation September–November 2024 (12 weeks).

**Results.** Post-improvement survey 1–31 December 2024, 300 opportunities observed. Non-compliance 15.3% (46/300). Improvement rate = (37.8% − 15.3%)/37.8% = 59.5%. Achievement rate = (37.8% − 15.3%)/(37.8% − 16%) = 103.2%. Target achieved. Non-compliance before patient contact fell from 38.2% to 12.0%, the largest improvement.

**Standardisation.** Revised the "General Medical Ward Hand-Hygiene Promotion Standard Operating Procedure" (SOP-IC-028 rev. 2) to include the replacement and refilling process for clip-on holders. Monthly audit results continue to be fed back immediately via LINE. New-staff training now includes a "five moments" scenario-recognition test. Intangible results: members report growth in the use of quality tools, team communication and problem analysis. Suggested next topic: reducing central-line-associated bloodstream infection.

**Withheld trap manifest.** Trap Q09 (topic selection; difficulty Medium): the topic-selection process does not present a comparison of candidate topics—a single topic goes straight to scoring, without the "several candidates → comparison matrix → highest priority" decision; the low compliance figure supports the topic but does not show it was the unit's top priority. Trap Q04 (countermeasures; difficulty Medium): countermeasure 1 (pocket card and quiz) corresponds weakly to true cause 1 (a knowledge–behaviour gap for the before-patient-contact moment); although countermeasure 3 provides immediate feedback for the programme as a whole, the specific behaviour gap lacks a bedside prompt, peer reminder, on-the-spot audit feedback or positive reinforcement, and the quiz tests knowledge rather than behaviour. Designed anchors: Q1 4, Q2 3, Q3 3, Q4 4, Q5 4, Q6 4, Q7 3, Q8 3.

#### Glossary of QCC terms as used in the benchmark

The Chinese terms appear in the case files of the data release; only English glosses are given here.

| Term | Meaning in the Taiwanese QCC convention |
|---|---|
| Quality Control Circle, QCC | Small front-line team that runs one improvement cycle on one topic and reports it in a fixed ten-step format. |
| Circle leader / facilitator | Member who leads the circle; senior staff member who coaches it. |
| Topic selection | Candidate topics scored on hospital policy, importance, urgency and circle capability; the highest total is chosen. |
| Current-state survey | Baseline measurement over a defined period, usually summarised in a Pareto chart with cumulative percentages. |
| Target-setting formula | Target = current value − current value × improvement focus (cumulative share of the Pareto items addressed) × circle capability (0–1). |
| Fishbone diagram | Cause-and-effect diagram with the categories man, machine, material, method, environment. |
| Candidate cause / true cause | Causes listed on the fishbone; the subset confirmed by verification (checklists, observation, data) as the causes to act on. |
| Countermeasure evaluation matrix | Each countermeasure scored on feasibility, effect and economy; adopted if above a threshold. |
| Effect confirmation | Post-implementation measurement over a period comparable to the baseline. |
| Improvement rate | (before − after)/before, for indicators that should decrease. |
| Achievement rate | (before − after)/(before − target). |
| Standardisation | Revision or creation of standard operating procedures, checklists and training so that the gain persists. |
| Intangible results radar chart | Self-rated before/after chart of team attributes (problem-solving, communication, cohesion, quality-tool skills, responsibility, enthusiasm). |

#### Crosswalk from the eight dimensions to QCC steps, rubric sources and trap types

| Dimension | Report fields the rubric reads | Trap types that degrade it (instances) | Note |
|---|---|---|---|
| Q1 Root Cause Depth | root_cause_analysis (depth of causal chain to system level) | none | assessed only through anchors |
| Q2 Systemic Coverage | root_cause_analysis (coverage of the five fishbone categories; rubric deducts one point per omitted category) | none by the case-format map | trap Q01 (fishbone category omitted) is mapped to Q5 by the specification although the rubric also penalises it here |
| Q3 Action Alignment | countermeasures (each countermeasure vs a verified cause; traceability) | Q04 (7) | |
| Q4 Data Foundation | current_status, target_setting, results (baseline completeness, Pareto, pre/post comparability) | Q05 (6), Q09 (5) | |
| Q5 Methodological Rigor | all steps (candidate topics, target formula, cause verification, PDCA completeness, formulas) | Q01 (6), Q02 (6), Q03 (8), Q04 (7), Q06 (5) | 32 of 65 trap-dimension pairs |
| Q6 Standardization | standardization, results (SOP/checklist concreteness, monitoring, formula correctness) | Q07 (5), Q08 (6) | |
| Q7 Evidence Traceability | root_cause_analysis, target_setting (data, literature or benchmark support) | none | assessed only through anchors |
| Q8 Innovation & Value | countermeasures, standardization (beyond training/SOP templates; plausibility of intangible results) | Q10 (4) | |

Trap types: Q01 fishbone main categories not covered; Q02 target set without a basis; Q03 cause verification missing; Q04 countermeasures not matched to verified causes; Q05 pre/post data not comparable; Q06 PDCA Check step missing; Q07 standardisation not established; Q08 achievement-rate formula wrong; Q09 topic selection without data support; Q10 intangible results exaggerated. The eight dimensions do not cover circle-process criteria used in competition scoring (team participation, activity planning, sustained effect after standardisation, dissemination to other units); Q6 combines standardisation with formula correctness.

### SM6. Supplementary File S11 – TRIPOD-LLM and GRRAS reporting checklists

#### TRIPOD-LLM

Items follow the fillable TRIPOD-LLM checklist (Supplementary Table 2 of the TRIPOD-LLM statement; research-design codes: E = LLM evaluation, H = evaluation in healthcare settings, M/D = LLM methods/de novo development). This study evaluates existing LLMs on a scoring task; items that apply only to LLM development, summarisation or patient/EHR data are marked not applicable.

| Item | Checklist item (abridged) | Where reported |
|---|---|---|
| 1 | Identify the study as developing, fine-tuning and/or evaluating an LLM; task, population, outcome | Title; Abstract |
| 2 | Abstract | Abstract (structured) |
| 3a | Healthcare context, rationale, existing approaches | Introduction |
| 3b | Target population, intended use in the care pathway, intended users | Introduction; Discussion (triage design, users: QCC reviewers and facilitators) |
| 4 | Objectives; development vs evaluation | Introduction (last paragraph) |
| 5a | Sources of data for evaluation and rationale | Methods (Study design); File S10 |
| 5b | Data points, distribution, languages, countries of origin | Methods (30 synthetic Traditional-Chinese cases, three quality levels, length; 8 PMC renderings); Table S6; File S10 |
| 5c | Dates of oldest and newest text in the evaluation data | Synthetic cases generated 1–6 May 2026; PMC source articles 2020–2025 (Table S6); scoring 15 September 2026 |
| 5d | Pre-processing and quality checking | Methods (Blinding); Supplementary Methods (generation, review of five cases, PMC rendering) |
| 5e | Missing and imbalanced data; reasons for omission | Methods (Missingness); File S8 |
| 6a | LLM name, version, last training date | Methods; File S7 (identifiers; training dates not disclosed by providers) |
| 6b | LLM development process | Not applicable (no development or fine-tuning) |
| 6c | Text generation, prompt engineering, inference settings | Methods (Evaluators and scoring: prompt v1.0, variants, temperature 0, 4,096-token limit, three runs); Supplementary Methods |
| 6d | Initial and post-processed output | Methods (YAML scores, evidence, defect list); Missingness (parsers); Supplementary Methods (defect parser) |
| 6e | Classification, probabilities, thresholds | Not applicable (ordinal scores; no probabilities) |
| 7a | Metrics for quality of generative outputs | Not applicable as generative-quality metrics; scoring reliability reported as ICC(A,1) (Methods) |
| 7b | Relevance of metrics to the downstream task; correlation with human evaluation | Discussion (reliability as precondition; no human-evaluation correlation available) |
| 7c | Outcome definition, how predictions were calculated, date of inference, metrics | Methods (Statistical analysis; 15 September 2026); File S7 |

| Item | Checklist item (abridged) | Where reported |
|---|---|---|
| 7d | Assessor qualifications, instructions, inter-assessor agreement for subjective outcomes | Methods (anchors emitted by the generator model, reviewed by the first author; no independent rater); Table S9 |
| 7e | Comparison with other LLMs, humans, benchmarks | Results (five evaluators; leave-one-out; $k$=5); no human comparison |
| 8a–c | Annotation process, number of annotators, agreement, annotator background | Supplementary Methods (generation; single-author review; no second annotator; no inter-annotator agreement available) |
| 9a | Prompt design, curation and selection | Methods; Supplementary Methods (v1.0 drafted from the case-format specification and protocol; v1.1/v1.2 tried informally, not adopted) |
| 9b | Data used to develop prompts | Supplementary Methods (case-format specification and the authors' earlier, unblinded pilot) |
| 10 | Summarisation pre-processing | Not applicable |
| 11 | Instruction tuning / alignment | Not applicable |
| 12 | Compute or cost | File S7 (570 API calls, USD 46.77) |
| 13 | Ethics approval / consent | Ethics statement (not required; synthetic and public data) |
| 14a | Funding and role of funders | Funding |
| 14b | Conflicts of interest | Conflicts of Interest |
| 14c | Protocol access | Sub-study protocol v0.3 (May 2026) available from the corresponding author; not deposited |
| 14d | Registration | Not registered |
| 14e | Data availability | Data Availability (upon reasonable request) |
| 14f | Code availability | Code Availability |
| 15 | Patient and public involvement | None |
| 16a–d | Flow and characteristics of patient/EHR data | Not applicable (no patient data) |
| 17 | Performance by pre-specified metrics | Results; Tables 1–3; Tables S1–S5, S9 |
| 18 | LLM updating | Not applicable |
| 19a | Overall interpretation, fairness | Discussion (first and last paragraphs) |
| 19b | Limitations, bias, uncertainty, generalisability | Discussion (limitations paragraph) |
| 19c | Challenges of the data for the domain (representation, missingness, bias) | Discussion (language and format, length confound, PMC rendering); File S8 |
| 19d | Intended use, input, end-user, level of autonomy | Discussion (triage design to be validated; human adjudication) |
| 19e | Handling of poor-quality or unavailable input | Discussion (safeguards); Methods (truncation, parse failures) |
| 19f | User interaction and expertise required | Discussion (facilitators; expert adjudication) |
| 19g | Next steps | Discussion (expert-labeled validation; length-matched benchmark) |

#### GRRAS

Items are summarised from the Guidelines for Reporting Reliability and Agreement Studies (Kottner et al., 2011; main-text reference list).

| Item | Item (summarised) | Where reported |
|---|---|---|
| 1 | Identify the study as a reliability or agreement study in title and abstract | Title (“reliability study”); Abstract |
| 2 | Rationale; which type of reliability or agreement is studied | Introduction; Methods (inter-model absolute agreement, ICC(A,1)) |
| 3 | Subjects, raters and the measurement instrument | Methods (30 cases; five LLM evaluators; scoring prompt and rubric); File S10 |
| 4 | Sampling of subjects and raters | Methods (synthetic cases by design at three quality levels; evaluators chosen as distinct model families) |
| 5 | Sample-size determination | Methods (protocol target: half-width 0.15 at ICC 0.75, $k$=4, 30–40 cases) |
| 6 | Selection and characteristics of subjects | Methods; Supplementary Methods (generation, quality levels, traps, length) |
| 7 | Rater characteristics, number, training | Methods (model identifiers, persona, prompt); File S7 |
| 8 | Blinding of raters | Methods (Blinding of evaluator inputs; evaluators independent of one another) |
| 9 | Order and timing of measurements | Methods (three independent runs per evaluator, 15 September 2026); Table S3 |
| 10 | Statistical analysis and coefficients used | Methods (ICC(A,1), McGraw–Wong intervals, cluster bootstrap, Cliff’s delta); Supplementary Methods |
| 11 | Number of raters, subjects and observations; missing data | Results; Table 1; File S8 |
| 12 | Estimates with confidence intervals | Results; Tables 1–2; Tables S1–S2 |
| 13 | Representativeness and generalisability | Discussion (limitations) |
| 14 | Comparison with other studies; practical relevance | Discussion |
| 15 | Availability of data and materials | Data and Code Availability (upon reasonable request) |
